# *MC1R* loss-of-function is associated with accelerated Parkinson’s disease motor decline

**DOI:** 10.64898/2025.12.26.25343003

**Authors:** Jackson G. Schumacher, Xinyuan Zhang, Jian Wang, Michael A. Schwarzschild, Marianna Cortese, Xiqun Chen

**Affiliations:** Department of Neurology, Massachusetts General Hospital and Harvard Medical School, Boston, MA 02129, USA; Aligning Science Across Parkinson’s (ASAP) Collaborative Research Network, Chevy Chase, MD 20815, USA; Channing Division of Network Medicine, Brigham and Women’s Hospital and Harvard Medical School, Boston, MA 02115, USA; Department of Biostatistics, The University of Texas MD Anderson Cancer Center, Houston, TX, 77030, USA; Department of Epidemiology, Harvard T.H. Chan School of Public Health, Boston, MA, 02115, USA; Department of Nutrition, Harvard T.H. Chan School of Public Health, Boston, MA, 02115, USA

**Author notes:** Denotes equal contribution (Jackson G. Schumacher, Xinyuan Zhang). Denotes corresponding author Correspondence to: Xiqun Chen, Department of Neurology, Massachusetts General Hospital and Harvard Medical School, Boston, MA 02129, USA.

## Abstract

**Background:** Melanocortin 1 receptor (*MC1R*) is a key regulator of pigmentation. Previous studies have linked *MC1R* loss-of-function variants to increased risk for Parkinson*’*s disease (PD); however, whether they are associated with PD progression remains unknown. Using data from the Parkinson*’*s Progression Markers Initiative (PPMI) cohort, we aimed to test whether *MC1R* loss-of-function variants, especially those previously associated with an increased risk of PD, are associated with PD progression and phenoconversion.

**Methods:** We analyzed PD progression in 802 PPMI participants (n=185 sporadic PD, 220 *MC1R* PD (139 heterozygotes, 64 compound heterozygotes, 17 homozygotes), 84 *LRRK2* PD, 43 *GBA* PD, 187 *MC1R + LRRK2* PD, and 83 *MC1R + GBA* PD) with 13 years of motor, non-motor, and cognitive assessments alongside 5 years of dopamine transporter imaging using linear mixed-effects models adjusted for potential confounders. Additionally, we explored risk of phenoconversion in 45 prodromal participants (n=16 sporadic, 29 *MC1R*) using time-to-event survival analysis.

**Results:** Participants with *MC1R* PD exhibited a 23% faster rate of motor decline (p=0.035) than participants with sporadic PD. The R160W variant (n=43 carriers), previously associated with an increased risk of PD, exhibited the strongest association with motor decline (p=0.023). High penetrance variants exhibited a stronger association with motor decline than low penetrance variants. Further, stratifying by genotype revealed that homozygotes exhibited a stronger association with motor decline than heterozygotes. Participants with *MC1R* PD also exhibited a non-statistically significant 24% faster rate of non-motor decline (p=0.070) than participants with sporadic PD. The rate of change in MoCA and DAT-SPECT SBR was minimal in all groups. Although not statistically significant, prodromal participants with *MC1R* loss-of-function variants exhibited a 3.81-fold increased risk of phenoconversion compared to noncarriers (p=0.059).

**Interpretation:** *MC1R* loss-of-function variants are associated with accelerated PD motor decline in the PPMI cohort. Together with previous biological findings from our group and others, these results highlight the potential role of MC1R in PD prognosis, warranting further validation and investigation into whether MC1R-targeted interventions may modify PD progression.

## Introduction

Parkinson*’*s disease (PD) is a progressive neurodegenerative disorder characterized by loss of dopaminergic neurons in the substantia nigra and α-synuclein pathology, manifesting clinically by motor symptoms such as tremor, rigidity, and bradykinesia, alongside cognitive and other non-motor impairments.^1^ While pathogenic mutations in *LRRK2* and *GBA* are known to influence disease susceptibility and clinical course, there are likely additional genetic factors contributing to variability in disease manifestation. Identifying these factors provides an opportunity to better understand the spectrum of disease progression and to inform more tailored clinical and research strategies.

Melanocortin 1 receptor (MC1R), best known for its role in skin and hair pigmentation, has been implicated in dopaminergic neuron vulnerability. *MC1R* loss-of-function variants, including red hair color (RHC) alleles, shift the tyrosine-levodopa-melanin synthesis pathway from eumelanin, a brown-black pigment with potent antioxidant and photoprotective properties, toward pheomelanin, a less stable, red-yellow pigment with intrinsic pro-oxidant potential.^2–4^ *MC1R* also modulates oxidative stress responses and cytoprotective pathways in other cell types, including dopaminergic neurons.^5^ Preclinical studies in *MC1R*-deficient mice show increased susceptibility of dopaminergic neurons to degeneration and motor deficits, supporting a potential role for MC1R in neuronal resilience.^6–9^

Population-based evidence further links MC1R and pigmentation traits to PD risk. Lighter skin and red hair, phenotypes associated with *MC1R* loss-of-function, have been reported to increase risk of PD.^10–13^ Moreover, melanoma, a malignancy strongly associated with *MC1R* variants, has been consistently linked to increased PD risk.^14^ Specific RHC alleles, including R151C and R160W, have been studied as potential PD risk variants, although findings remain inconsistent.^13,15–20^ Despite the convergence of preclinical and population-based evidence, whether *MC1R* RHC variants are associated with PD progression and phenoconversion from prodromal states to clinically diagnosed PD remains unknown.

To address this knowledge gap, we leveraged detailed longitudinal data from the Parkinson*’*s Progression Markers Initiative (PPMI) to evaluate whether *MC1R* RHC variants, including penetrance-defined subgroups, heterozygotes, and homozygotes, and individual variants, are associated with PD progression and phenoconversion.

## Methods

### Analysis design and participants

This longitudinal cohort analysis utilizes data from PPMI and includes participants from the Parkinson*’*s Disease Cohort, Genetic Registry, and Prodromal Cohort. Genome-wide sequencing data from PPMI was used to categorize participants with PD into six groups: participants with sporadic PD (non-carriers of any of the following variants), participants with *MC1R* PD (carriers of *MC1R* variants R163Q/P, V60L, V92M/L, R151S/G/C, R160W, D294N/H, D84E, or R142H variants), participants with *LRRK2* PD (carriers of *LRRK2* variants G2019S, R1441C/G + M1646T, or N2081D/N14D variants), participants with *GBA* PD (carriers of *GBA* variants E326K, R502C, A495P, or N409S variants), participants with *MC1R + LRRK2* PD (carriers of any combination of the aforementioned *MC1R* and *LRRK2* variants), and participants with *MC1R + GBA* PD (carriers of any combination of the aforementioned *MC1R* and *GBA* variants).

Participants with *MC1R* PD were further stratified by penetrance, genotype, and variant. In subanalyses stratifying participants with *MC1R* PD by penetrance and variant, compound heterozygote participants were included in multiple groups. PD participants who met enrollment criteria but had scans without evidence of dopamine deficiency (n=66) were excluded. Additionally, PD participants carrying any *SNCA* (n=38), *PRKN* (n=9), *PINK1* (n=1), *PARK7* (n=1), or *VPS35* (n=1) variants, as well as those carrying both *LRRK2* and *GBA* variants (n=24), were excluded. Participants with prodromal PD were categorized as either sporadic or *MC1R* prodromal PD. Prodromal participants with *LRRK2* (n=60), *GBA* (n=41), *SNCA* (n=2), *PRKN* (n=1), *PARK7* (n=1), or *VPS35* (n=1) variants were excluded.

The data analyzed was collected by PPMI between the study*’*s inception, July 7, 2010, and October 1, 2025. All publicly available data at the time of retrieval were analyzed. More detailed information about PPMI*’*s mission and inclusion criteria, as well as sources and methods of participant selection, can be found on the PPMI website (www.ppmi-info.org/access-data-specimens/download-data). The data was last accessed using the PPMI portal on October 15, 2025.

### Clinical data

Participants underwent clinical assessments and imaging by the PPMI team upon recruitment into the cohort and during subsequent follow-up visits as previously described.^21,22^ We analyzed 13 years (mean follow-up = 5.33 years) of Movement Disorder Society Unified Parkinson*’*s Disease Rating Scale Part III (MDS-UPDRS III), Part I (MDS-UPDRS I), and Montreal Cognitive Assessment (MoCA) data as well as 5 years (mean follow-up = 2.86 years) of dopamine transporter (DAT) imaging with single-photon emission computed tomography (DAT-SPECT) as measures of PD-related motor decline, non-motor decline, cognitive decline, and loss of DAT.^23–25^ Higher MDS-UPDRS III and I scores reflect more severe motor and non-motor features, while lower MoCA scores reflect greater cognitive impairment. Only *“*off*”* medication state MDS-UPDRS III assessments were analyzed to minimize the influence of dopaminergic medications on motor scores.

### Statistics

All PPMI participants with genome-wide sequencing data and a diagnosis of PD or prodromal PD at the time of data retrieval were included in our analyses. A linear mixed-effects model with years since baseline as the time scale was used to estimate the yearly rate of change in MDS-UPDRS III, the primary outcome measure, as well as MDS-UPDRS I, MoCA, and DAT-SPECT specific binding ratio (SBR). The primary exposure was genetic group, and each outcome was modeled independently. To estimate group-specific progression rates in each outcome measure, we included a genetic group-by-time interaction term. To ensure unbiased subgroup comparisons, separate models were used when stratifying by sex, penetrance, genotype, and variant. All models included participant-level random intercepts and slopes for time with an unstructured covariance matrix and allowed for heteroscedastic variance between groups. Based on a review of existing literature, all models were adjusted for age, sex, race, ethnicity, time from original diagnosis to baseline, and assessment score at baseline.^26–28^ To better control for the effect of PD medications, the MDS-UPDRS III model was adjusted for levodopa equivalent daily dosage (LEDD) at the time of each visit. A significance level of α=0.05 was used for all analyses. Given the exploratory nature of these analyses, formal adjustment for multiple comparisons was not performed. All progression analyses and associated figures were generated using R software version 4.4.1.

Risk of phenoconversion from prodromal PD to clinically diagnosed PD was analyzed using a Cox proportional hazards model adjusted for participants*’* demographic characteristics. All phenoconversion analysis and associated figures were generated using SAS® software version 9.4.

### Role of the funding source

The funders had no role in the study design, analysis, interpretation of data, or the writing of the report. While the funders were not directly involved in data collection, PPMI is funded by the Michael J. Fox Foundation and is associated with the Aligning Science Across Parkinson*’*s initiative.

## Results

### Baseline demographics and clinical characteristics for PD participants

Here, we analyzed longitudinal data from 802 PD participants in the PPMI cohort, including 185 with sporadic PD, 220 with *MC1R* PD, 84 with *LRRK2* PD, 43 with *GBA* PD, 187 with *MC1R + LRRK2* PD, and 83 with *MC1R + GBA* PD. Baseline demographic and clinical characteristics for each group are displayed in Table 1. No statistical analysis on demographic characteristics between groups was performed; however, participants with *MC1R* PD were younger at disease onset (61.5 vs. 65.2), more likely to be male (66.8% vs. 60.5%), and more likely to have a history of melanoma (2.25% vs. 0.54%), compared to those with sporadic PD (Table 1).

**Table 1.** Baseline demographic and clinical characteristics for sporadic, *MC1R, LRRK2, GBA, MC1R + LRRK2*, and *MC1R + GBA* PD. Data is shown as n (%), %, mean (standard deviation), or median (interquartile range). Statistical analysis comparing group characteristics was not performed. Abbreviations: PD, Parkinson*’*s disease; *MC1R*, melanocortin 1 receptor; *LRRK2*, leucine-rich repeat kinase 2; GBA, glucocerebrosidase; SAA, seed amplification assay.

|  | Sporadic PD | MC1R PD | LRRK2 PD | GBA PD | MC1R + LRRK2 PD | MC1R + GBA PD |
| --- | --- | --- | --- | --- | --- | --- |
| N | 185 | 220 | 84 | 43 | 187 | 83 |
| MC1R compound heterozygotes | 0 | 64 (29.1) | 0 | 0 | 46 (24.6) | 17 (20.5) |
| MC1R homozygotes | 0 | 17 (7.73) | 0 | 0 | 16 (8.55) | 6 (7.23) |
| Age at baseline (years) | 65.2 (9.88) | 61.5 (10.2) | 63.9 (10.1) | 61.1 (10.9) | 66.7 (10.3) | 64.2 (9.21) |
| Years since original diagnosis | 0.76 (1.51) | 0.62 (0.75) | 1.79 (2.00) | 1.53 (2.12) | 1.65 (2.12) | 1.70 (2.19) |
| Age at PD onset (years) | 64.4 (9.96) | 60.8 (10.2) | 62.1 (10.8) | 59.6 (11.4) | 65.1 (10.9) | 62.5 (9.75) |
| Male | 112 (60.5) | 147 (66.8) | 48 (57.1) | 22 (51.2) | 87 (46.5) | 52 (62.7) |
| Race |  |  |  |  |  |  |
| White | 168 (90.8) | 199 (90.5) | 68 (81.0) | 41 (95.3) | 175 (93.6) | 82 (98.8) |
| Asian | 1 (0.54) | 7 (3.17) | 1 (1.20) | 0 | 0 | 0 |
| Black | 4 (2.16) | 3 (1.35) | 0 | 0 | 0 | 0 |
| Multiracial | 7 (3.78) | 6 (2.73) | 11 (13.0) | 2 (4.70) | 11 (5.88) | 0 |
| Unknown | 5 (2.70) | 5 (2.25) | 4 (4.80) | 0 | 0 | 1 (1.20) |
| Hispanic or Latino ethnicity | 19 (10.3) | 4 (1.82) | 26 (30.9) | 0 | 23 (12.3) | 3 (3.61) |
| Relatives with PD |  |  |  |  |  |  |
| Parent | 43 (23.2) | 21 (9.55) | 30 (35.7) | 6 (14.0) | 72 (38.5) | 16 (19.3) |
| Other | 46 (24.9) | 39 (17.7) | 33 (39.3) | 12 (27.9) | 84 (44.9) | 19 (22.9) |
| α-Synuclein SAA status |  |  |  |  |  |  |
| Positive | 112 | 203 | 41 | 20 | 70 | 46 |
| Negative | 9 | 8 | 12 | 0 | 39 | 4 |
| Data not available | 64 | 9 | 31 | 23 | 78 | 33 |
| α-Synuclein SAA positivity | 92.6 | 96.2 | 77.4 | 100 | 64.2 | 92.0 |
| History of melanoma | 1 (0.54) | 5 (2.25) | 0 | 0 | 1 (0.53) | 1 (1.20) |
| Clinical Assessments |  |  |  |  |  |  |
| Average follow-up (years) | 6.15 (4.04) | 6.84 (3.77) | 4.58 (3.17) | 3.96 (4.18) | 3.96 (3.64) | 4.10 (3.38) |
| MDS-UPDRS I score at baseline | 4.41 (3.18) | 4.62 (3.44) | 4.98 (3.97) | 4.32 (3.17) | 6.25 (4.56) | 5.89 (4.07) |
| MDS-UPDRS III score at baseline | 22.7 (9.6) | 20.2 (9.04) | 18.3 (10.3) | 25.9 (15.4) | 20.3 (10.4) | 24.4 (10.8) |
| MoCA score at baseline | 26.8 (2.35) | 27.1 (2.58) | 25.9 (2.88) | 26.8 (3.19) | 25.9 (3.12) | 26.5 (2.57) |
| DAT-SPECT SBR at baseline |  |  |  |  |  |  |
| Average follow-up (years) | 3.20 (1.45) | 3.08 (1.40) | 3.00 (1.41) | 2.86 (1.49) | 2.36 (1.70) | 2.51 (1.62) |
| Caudate | 1.93 (1.63, 2.28) | 1.92 (1.62, 2.35) | 2.07 (1.61, 2.31) | 2.04 (1.39, 2.65) | 1.81 (1.41, 2.16) | 1.69 (1.34, 2.12) |
| Putamen | 0.77 (0.63, 0.91) | 0.81 (0.65, 1.00) | 0.78 (0.58, 0.99) | 0.81 (0.63, 0.89) | 0.67 (0.56, 0.87) | 0.59 (0.53, 0.82) |

Out of the 802 PD participants analyzed, 490 (61.1%) carried at least one *MC1R* RHC variant, including 324 (40.4%) who were heterozygous for a single variant, 127 (15.8%) who were heterozygous for two variants, and 39 (4.86%) who were homozygous for a single variant. The minor allele frequency of *MC1R* RHC variants in the PPMI cohort compared to other PD cohorts is displayed in Table 2. Baseline demographic and clinical characteristics for participants with *MC1R* PD stratified by penetrance, genotype, and variant are displayed in Supplemental Tables 1, 2, and 3.

**Table 2.** *MC1R* red hair variant minor allele frequency in the PPMI cohort compared to other PD cohorts. Minor allele frequency was calculated by 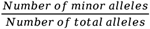. Number of total alleles = 802 x 2 = 1604. R = high penetrance, r = low penetrance. Abbreviations: MAF, minor allele frequency.

|  | Penetrance | PPMI<br>(N=802) | Lorenzo-Betancor et al.<br>2016<br>(N=889) | Gan-Or et al.<br>2016 (SPOT)<br>(N=539) | Gan-Or et al. 2016<br>(MNI)<br>(N=551) | Gao et al. 2009<br>(HPFS/NHS)<br>(N=298) | Dong et al.<br>2014 (PAGE)<br>(N=808) | Tell-Marti et<br>al. 2015<br>(N=870) | Lubbe et al.<br>2016 (NeuroX)<br>(N=5,944) |
| --- | --- | --- | --- | --- | --- | --- | --- | --- | --- |
| All variants | ---- | 0.439 | 0.469 | ---- | ---- | ---- | ---- | ---- | ---- |
| r variants | ---- | 0.297 | 0.267 | 0.329 | 0.232 | ---- | ---- | ---- | ---- |
| R variants | ---- | 0.143 | 0.202 | ---- | ---- | ---- | ---- | ---- | ---- |
| R163Q/P (rs885479) | r | 0.049 | 0.049 | 0.064 | 0.024 | ---- | ---- | ---- | ---- |
| V60L (rs1805005) | r | 0.163 | 0.129 | 0.195 | 0.142 | ---- | ---- | ---- | ---- |
| V92M/L (rs2228479) | r | 0.064 | 0.089 | 0.070 | 0.066 | ---- | ---- | ---- | ---- |
| R151S/G/C (rs1805007) | R | 0.056 | 0.085 | 0.065 | 0.054 | 0.091 | ---- | ---- | ---- |
| R160W (rs1805008) | R | 0.053 | 0.079 | 0.045 | 0.048 | ---- | 0.069 | 0.025 | 0.064 |
| D294N/H (rs1805009) | R | 0.008 | 0.018 | 0.011 | 0.018 | ---- | ---- | ---- | ---- |
| D84E (rs1805006) | R | 0.006 | 0.011 | ---- | ---- | ---- | ---- | ---- | ---- |
| R142H (rs11547464) | R | 0.009 | 0.009 | ---- | ---- | ---- | ---- | ---- | ---- |

### *MC1R* RHC variants are associated with accelerated motor decline

The rate of change in MDS-UPDRS III score for participants with sporadic PD (n=185) was 1.98 (95% confidence interval: 1.64 to 2.32) points per year (Figure 1 and Table 3). Participants with *MC1R* PD (n=220) had a higher rate of change in MDS-UPDRS III than those with sporadic PD (2.44; β diff=0.46 (0.03 to 0.88) p=0.035)(Figure 1 and Table 3). Given the possible influence of MC1R on the metabolism of exogenous levodopa, we performed the same analysis without adjusting for LEDD and found similar results (Table 3). When stratifying by sex, although the rate of change in MDS-UPDRS III was numerically different between males with *MC1R* PD (2.49; β diff=0.44 (−0.02 to 0.90) p=0.063) and females with *MC1R* PD (2.24; β diff=0.44 (−0.43 to 1.31) p=0.324), the effect of *MC1R* relative to sporadic PD appeared similar across sexes (Table 3).

**Table 3.** Comparison of the rate of change in MDS-UPDRS III between participants with sporadic and *MC1R* PD. Slopes (unit per year) were estimated from linear mixed models. Fixed effects included time ^*^ genetic group, baseline MDS-UPDRS III, baseline age, years since original diagnosis at baseline, sex, race, ethnicity, and LEDD. Participant-level random intercepts and slopes with unstructured covariance, heteroscedastic by genetic group. #1–5 denotes the model as stratification by sex, penetrance, genotype, and variant were performed in separate models using sporadic PD as the reference group. N refers to the comparison group | reference group. A single participant may be included in several groups due to the presence of compound heterozygotes. Abbreviations: PD, Parkinson*’*s disease; *MC1R*, melanocortin 1 receptor; *LRRK2*, leucine-rich repeat kinase 2; GBA, glucocerebrosidase; MDS-UPDRS III, Movement Disorder Society Unified Parkinson*’*s Disease Rating Scale Part III

| Group | N | MDS-UPDRS III rate of change<br>(with LEDD adj.) |  |  | MDS-UPDRS III rate of change<br>(without LEDD adj.) |  |  |
| --- | --- | --- | --- | --- | --- | --- | --- |
| | | Slope, $\beta$ (95% CI) | % diff | p-value | Slope, $\beta$ (95% CI) | % diff | P-value |
| Sporadic PD <sup>#1</sup> | 185 | 1.98 (1.64, 2.32) |  |  | 1.80 (1.47, 2.13) |  |  |
| Male Sporadic PD <sup>#2</sup> | 112 | 2.05 (1.67, 2.43) |  |  | 1.86 (1.50, 2.22) |  |  |
| Female Sporadic PD <sup>#2</sup> | 73 | 1.80 (1.12, 2.48) |  |  | 1.71 (1.04, 2.38) |  |  |
| *MC1R* PD vs. Sporadic PD <sup>#1</sup> | 220 \| 185 | 2.44, 0.46 (0.03, 0.88) | 23% | **0.035** | 2.25, 0.45 (0.03, 0.87) | 25% | **0.036** |
| Male *MC1R* PD <sup>#2</sup> | 147 \| 185 | 2.49, 0.44 (-0.02, 0.90) | 21% | 0.063 | 2.31, 0.45 (-0.01, 0.90) | 24% | 0.058 |
| Female *MC1R* PD <sup>#2</sup> | 73 \| 185 | 2.24, 0.44 (-0.43, 1.31) | 24% | 0.324 | 2.14, 0.43 (-0.45, 1.30) | 25% | 0.339 |
| r variant *MC1R* PD <sup>#3</sup> | 150 \| 185 | 2.32, 0.34 (-0.16, 0.83) | 17% | 0.183 | 2.13, 0.33 (-0.16, 0.82) | 18% | 0.185 |
| R variant *MC1R* PD <sup>#3</sup> | 102 \| 185 | 2.57, 0.59 (0.08, 1.11) | 30% | **0.025** | 2.38, 0.58 (0.07, 1.10) | 32% | **0.025** |
| Single Heterozygote *MC1R* PD <sup>#4</sup> | 139 \| 185 | 2.42, 0.44 (-0.03, 0.91) | 22% | 0.069 | 2.24, 0.44 (-0.03, 0.91) | 24% | 0.066 |
| Compound Heterozygote *MC1R* PD <sup>#4</sup> | 64 \| 185 | 2.37, 0.39 (-0.20, 0.98) | 20% | 0.197 | 2.17, 0.37 (-0.21, 0.96) | 21% | 0.215 |
| Homozygote *MC1R* PD <sup>#4</sup> | 17 \| 185 | 2.97, 0.99 (-0.20, 2.17) | 50% | 0.102 | 2.78, 0.98 (-0.20, 2.16) | 54% | 0.104 |
| R163Q/P PD <sup>#5</sup> | 41 \| 185 | 2.32, 0.34 (-0.32, 1.00) | 17% | 0.317 | 2.15, 0.35 (-0.31, 1.01) | 19% | 0.295 |
| V60L PD <sup>#5</sup> | 77 \| 185 | 2.22, 0.24 (-0.29, 0.77) | 12% | 0.376 | 2.04, 0.24 (-0.29, 0.77) | 13% | 0.377 |
| V92M/L PD <sup>#5</sup> | 54 \| 185 | 2.57, 0.59 (-0.18, 1.37) | 30% | 0.135 | 2.36, 0.56 (-0.22, 1.33) | 31% | 0.159 |
| R151S/G/C PD <sup>#5</sup> | 50 \| 185 | 2.49, 0.51 (-0.18, 1.19) | 26% | 0.147 | 2.30, 0.50 (-0.19, 1.18) | 28% | 0.156 |
| R160W PD <sup>#5</sup> | 43 \| 185 | 2.86, 0.88 (0.12, 1.64) | 44% | **0.023** | 2.68, 0.88 (0.12, 1.64) | 49% | **0.023** |
| D294N/H PD <sup>#5</sup> | 11 \| 185 | 3.34, 1.36 (-0.04, 2.76) | 69% | 0.057 | 3.17, 1.37 (-0.03, 2.77) | 76% | 0.054 |
| D84E PD <sup>#5</sup> | 4 \| 185 | 1.23, -0.75 (-2.99, 1.50) | -38% | 0.515 | 1.11, -0.69 (-2.93, 1.54) | -38% | 0.543 |
| R142H PD <sup>#5</sup> | 3 \| 185 | 2.61, 0.63 (-1.59, 2.84) | 32% | 0.580 | 2.48, 0.68 (-1.53, 2.89) | 38% | 0.545 |

**Figure 1.**
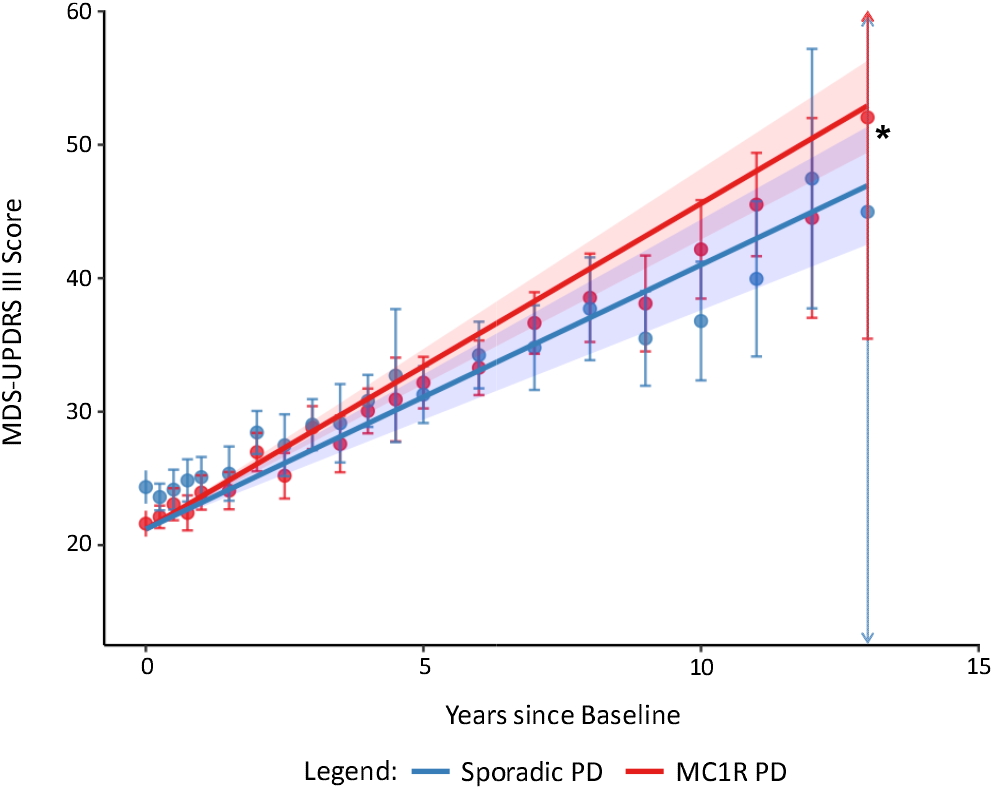
Comparison of the rate of change in MDS-UPDRS III between participants with sporadic and *MC1R* PD. Observed and predicted change in MDS-UPDRS III adjusted for age, time since diagnosis, sex, race, ethnicity, years of education, levodopa equivalent daily dosage, and baseline score in participants with sporadic PD (n=185; 1.98 (95% confidence interval: 1.64 to 2.32) and MC1R PD (n=220; 2.44; β diff=0.46 (0.03 to 0.88) p=0.035). Comparing the difference in change to participants with sporadic PD. Data points represent the mean observed value at the corresponding time point. Lines represent the predicted value over time after adjustment. Error bars and shading represent 95% confidence intervals for observed and predicted values. Arrows represent that the error bar extends beyond the range of the graph. ^*^p<0.05 Abbreviations: PD, Parkinson*’*s disease; MC1R, melano ortin 1 receptor, MDS-UPDRS III, Movement Disorder Society Unified Parkinson*’*s Disease Rating Scale Part III

When stratifying participants with *MC1R* PD by penetrance, both participants with low penetrance r variants (n=150)(2.32; β diff=0.34 (−0.16 to 0.83) p=0.183) and participants with high penetrance R variants (n=102)(2.57; β diff=0.59 (0.08 to 1.11) p=0.025) had a higher rate of change in MDS-UPDRS III compared to participants with sporadic PD; however, only the difference for participants with high penetrance R variants reached statistical significance (Table 3).

We also stratified participants with *MC1R* PD by genotype and found that although statistical significance was lost, single heterozygotes (n=139)(2.42; β diff=0.44 (−0.03 to 0.91) p=0.069), compound heterozygotes (n=64)(2.37; β diff=0.39 (−0.20 to 0.98) p=0.197), and homozygotes (n=17)(2.97; β diff=0.99 (−0.20 to 2.17) p=0.102) all had a nominally higher rate of change in MDS-UPDRS III compared to participants with sporadic PD (Table 3).

Finally, variant-specific subanalyses revealed that participants carrying the R160W (n=43) variant, which has previously been studied as a risk factor for PD, exhibited a statistically significant higher rate of change in MDS-UPDRS III compared to non-carriers with sporadic PD (2.86; β diff=0.88 (0.12 to 1.64) p=0.023). Similarly, the R151C variant has also been linked to PD risk, and we found that R151S/G/C (n=50) carriers exhibited a nominally higher rate of change in MDS-UPDRS III than non-carriers with sporadic PD (2.49; β diff=0.51 (−0.18 to 1.19) p=0.147). All other variants except for D84E were also associated with a nominally higher rate of change in MDS-UPDRS III (Table 3).

To assess whether *MC1R* variants modify the association between known PD-related genetic variants and motor progression, we examined MDS-UPDRS III trajectories in participants with *LRRK2* and *GBA* mutations. Participants with *MC1R* + *LRRK2* PD (n=187; 2.14; β diff=0.60 (−0.11 to 1.32) p=0.098) exhibited a nominally higher rate of change in MDS-UPDRS III than participants with *LRRK2* PD (n=84; 1.54 (0.93 to 2.15)) while participants with *MC1R* + *GBA* PD (n=83; 2.39, β diff=-0.60 (−1.92, 0.71) p=0.366) exhibited a nominally lower rate of change in MDS-UPDR III than those with *GBA* PD (n=43; 2.99 (1.90 to 4.09))(Table 4). None of the differences for participants with *LRRK2, MC1R* + *LRRK2, GBA*, or *MC1R* + *GBA* PD reached statistical significance.

**Table 4.**
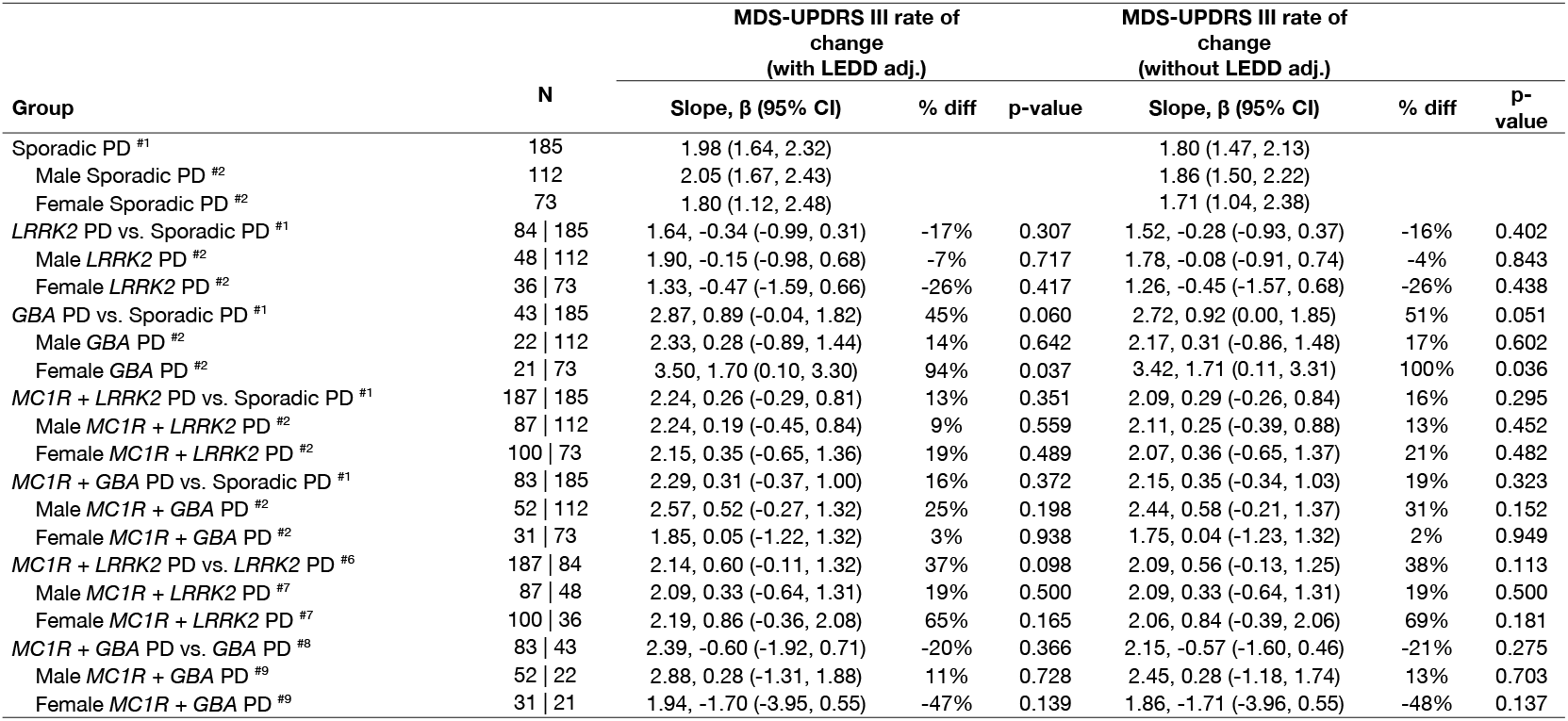
Comparison of the rate of change in MDS-UPDRS III between sporadic, *LRRK2, GBA, MC1R + LRRK2*, and *MC1R + GBA* PD. Slopes (unit per year) were estimated from linear mixed models. Fixed effects included time ^*^ genetic group, baseline MDS-UPDRS III, baseline age, years since original diagnosis at baseline, race, ethnicity, and LEDD. Participant-level random intercepts and slopes with unstructured covariance, heteroscedastic by genetic group. #1–7 denotes the model as *MC1R* PD, *LRRK2* PD, and *GBA* PD were compared to sporadic PD in a single model while *MC1R + LRRK2* PD and *MC1R + GBA* PD were compared to *LRRK2* PD and *GBA* PD in separate models. Further, stratification by sex was performed in a separate model using sporadic PD, *LRRK2* PD, and *GBA* PD as the reference groups where applicable. N refers to the comparison group | reference group. Abbreviations: PD, Parkinson*’*s disease; *MC1R*, melanocortin 1 receptor; *LRRK2*, leucine-rich repeat kinase 2; GBA, glucocerebrosidase; MDS-UPDRS III, Movement Disorder Society Unified Parkinson*’*s Disease Rating Scale Part III

| Group | N | MDS-UPDRS III rate of change<br>(with LEDD adj.) |  |  | MDS-UPDRS III rate of change<br>(without LEDD adj.) |  |  |
| --- | --- | --- | --- | --- | --- | --- | --- |
| | | Slope, $\beta$ (95% CI) | % diff | p-value | Slope, $\beta$ (95% CI) | % diff | P-value |
| Sporadic PD <sup>#1</sup> | 185 | 1.98 (1.64, 2.32) |  |  | 1.80 (1.47, 2.13) |  |  |
| Male Sporadic PD <sup>#2</sup> | 112 | 2.05 (1.67, 2.43) |  |  | 1.86 (1.50, 2.22) |  |  |
| Female Sporadic PD <sup>#2</sup> | 73 | 1.80 (1.12, 2.48) |  |  | 1.71 (1.04, 2.38) |  |  |
| *LRRK2* PD vs. Sporadic PD <sup>#1</sup> | 84 \| 185 | 1.64, -0.34 (-0.99, 0.31) | -17% | 0.307 | 1.52, -0.28 (-0.93, 0.37) | -16% | 0.402 |
| Male *LRRK2* PD <sup>#2</sup> | 48 \| 112 | 1.90, -0.15 (-0.98, 0.68) | -7% | 0.717 | 1.78, -0.08 (-0.91, 0.74) | -4% | 0.843 |
| Female *LRRK2* PD <sup>#2</sup> | 36 \| 73 | 1.33, -0.47 (-1.59, 0.66) | -26% | 0.417 | 1.26, -0.45 (-1.57, 0.68) | -26% | 0.438 |
| *GBA* PD vs. Sporadic PD <sup>#1</sup> | 43 \| 185 | 2.87, 0.89 (-0.04, 1.82) | 45% | 0.060 | 2.72, 0.92 (0.00, 1.85) | 51% | 0.051 |
| Male *GBA* PD <sup>#2</sup> | 22 \| 112 | 2.33, 0.28 (-0.89, 1.44) | 14% | 0.642 | 2.17, 0.31 (-0.86, 1.48) | 17% | 0.602 |
| Female *GBA* PD <sup>#2</sup> | 21 \| 73 | 3.50, 1.70 (0.10, 3.30) | 94% | 0.037 | 3.42, 1.71 (0.11, 3.31) | 100% | 0.036 |
| *MC1R* + *LRRK2* PD vs. Sporadic PD <sup>#1</sup> | 187 \| 185 | 2.24, 0.26 (-0.29, 0.81) | 13% | 0.351 | 2.09, 0.29 (-0.26, 0.84) | 16% | 0.295 |
| Male *MC1R* + *LRRK2* PD <sup>#2</sup> | 87 \| 112 | 2.24, 0.19 (-0.45, 0.84) | 9% | 0.559 | 2.11, 0.25 (-0.39, 0.88) | 13% | 0.452 |
| Female *MC1R* + *LRRK2* PD <sup>#2</sup> | 100 \| 73 | 2.15, 0.35 (-0.65, 1.36) | 19% | 0.489 | 2.07, 0.36 (-0.65, 1.37) | 21% | 0.482 |
| *MC1R* + *GBA* PD vs. Sporadic PD <sup>#1</sup> | 83 \| 185 | 2.29, 0.31 (-0.37, 1.00) | 16% | 0.372 | 2.15, 0.35 (-0.34, 1.03) | 19% | 0.323 |
| Male *MC1R* + *GBA* PD <sup>#2</sup> | 52 \| 112 | 2.57, 0.52 (-0.27, 1.32) | 25% | 0.198 | 2.44, 0.58 (-0.21, 1.37) | 31% | 0.152 |
| Female *MC1R* + *GBA* PD <sup>#2</sup> | 31 \| 73 | 1.85, 0.05 (-1.22, 1.32) | 3% | 0.938 | 1.75, 0.04 (-1.23, 1.32) | 2% | 0.949 |
| *MC1R* + *LRRK2* PD vs. *LRRK2* PD <sup>#6</sup> | 187 \| 84 | 2.14, 0.60 (-0.11, 1.32) | 37% | 0.098 | 2.09, 0.56 (-0.13, 1.25) | 38% | 0.113 |
| Male *MC1R* + *LRRK2* PD <sup>#7</sup> | 87 \| 48 | 2.09, 0.33 (-0.64, 1.31) | 19% | 0.500 | 2.09, 0.33 (-0.64, 1.31) | 19% | 0.500 |
| Female *MC1R* + *LRRK2* PD <sup>#7</sup> | 100 \| 36 | 2.19, 0.86 (-0.36, 2.08) | 65% | 0.165 | 2.06, 0.84 (-0.39, 2.06) | 69% | 0.181 |
| *MC1R* + *GBA* PD vs. *GBA* PD <sup>#8</sup> | 83 \| 43 | 2.39, -0.60 (-1.92, 0.71) | -20% | 0.366 | 2.15, -0.57 (-1.60, 0.46) | -21% | 0.275 |
| Male *MC1R* + *GBA* PD <sup>#9</sup> | 52 \| 22 | 2.88, 0.28 (-1.31, 1.88) | 11% | 0.728 | 2.45, 0.28 (-1.18, 1.74) | 13% | 0.703 |
| Female *MC1R* + *GBA* PD <sup>#9</sup> | 31 \| 21 | 1.94, -1.70 (-3.95, 0.55) | -47% | 0.139 | 1.86, -1.71 (-3.96, 0.55) | -48% | 0.137 |

When we examined the relationship between genetic status and rate of change in non-motor decline, we found that participants with *MC1R* PD (0.52; β diff=0.10 (−0.01 to 0.20) p=0.070) exhibited a nominally higher rate of change in MDS-UPDRS I than participants with sporadic PD (0.42 (0.34 to 0.50)). This trend persisted when stratifying participants with *MC1R* PD by penetrance, genotype, and variant (Supplemental Table 4). Associations between genetic status and the rate of change in MoCA and DAT-SPECT SBR were limited in all groups (Supplemental Table 4)

### Baseline demographics and risk of phenoconversion for prodromal participants

We explored the risk of phenoconversion to clinically diagnosed PD in 45 (n=16 sporadic and 29 *MC1R*) participants with prodromal PD. Baseline demographic characteristics for each group are displayed in Supplemental Table 5. At the time of data extraction, 22 participants had phenoconverted to PD (n=4 sporadic prodromal PD, 18 *MC1R* prodromal PD). Although not statistically significant, participants with *MC1R* prodromal PD exhibited a notable trend toward higher risk of phenoconversion compared with sporadic prodromal participants (Hazard ratio: 4.81 (0.94 to 24.5), p=0.059)(Figure 2).

**Figure 2.**
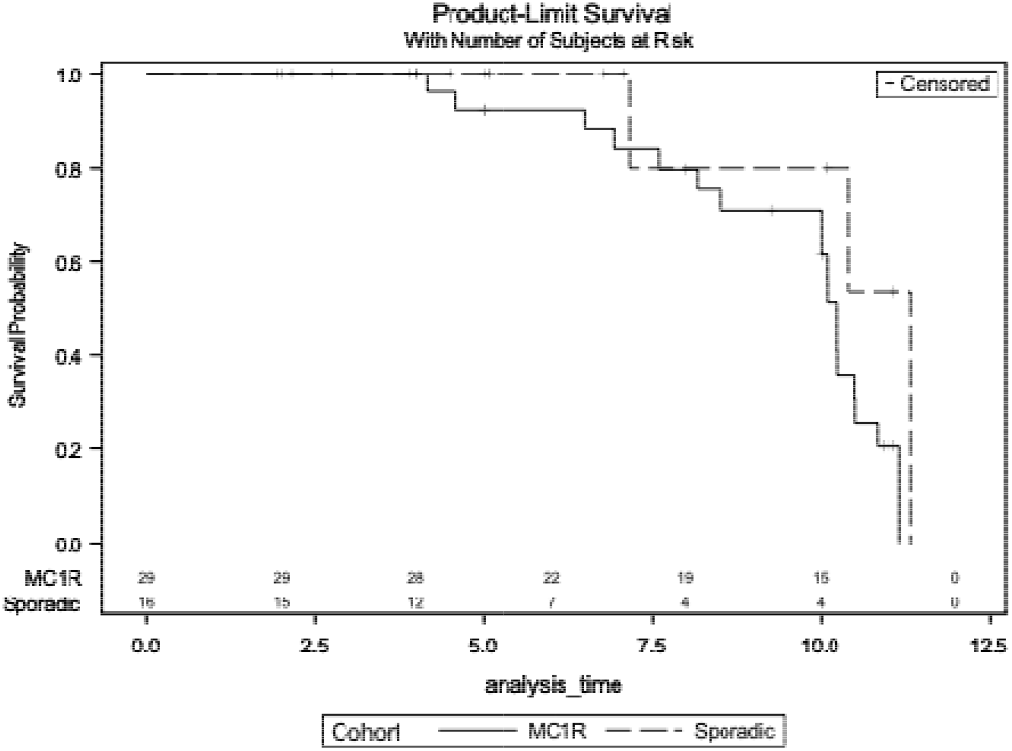
Risk of phenoconversion to clinically diagnosed PD in participants with sporadic and MC1R prodromal PD. The risk of phenoconversion in participants with sporadic and MC1R prodromal PD as assessed by Cox proportional hazards model adjusted for demographic characteristics. Hazard ratio: 4.81 (0.94 to 24.5) p=0.059. Abbreviations: PD, Parkinson*’*s disease; MC1R, melanocortin 1 receptor,

## Discussion

Using longitudinal data from the PPMI cohort, we found that *MC1R* RHC variants are associated with accelerated PD motor decline. High penetrance variants, R160W in particular, have the strongest associations. Although statistical significance was lost, all other variants except for D84E exhibited a higher rate of motor decline. Notably, *MC1R* appeared to modify motor decline even in participants carrying both an *MC1R* and *LRRK2* variant, as a trend towards slower motor decline in participants with *LRRK2* PD was not observed in participants with *MC1R* + *LRRK2* PD. Although statistical significance was not reached, prodromal participants with *MC1R* loss-of-function variants exhibited an increased risk of phenoconversion.

Previous studies examining the risk of PD in individuals with *MC1R* RHC variants have reported inconsistent results, with some finding an association between R151C, R160W, and PD risk, while others have not.^13,15–20^ Our finding that *MC1R* RHC variants are associated with accelerated motor decline indicates a role in disease course, even as evidence for their association with PD risk remains uncertain. Lighter skin and hair color have also been linked to an increased risk of PD. Individuals of European descent are two times more likely to be diagnosed with PD than individuals of African descent after adjustment for socioeconomic factors.^11^ Similarly, individuals with red hair were found to have a twofold higher risk for PD than individuals with black hair; however, that was not confirmed in a follow-up study on older participants in the same cohorts.^12,13^ Although *MC1R* loss-of-function heterozygotes do not typically have red hair, they may still have increased pheomelanin production and melanoma risk.^2–4,29^ In line with this, we observed a non-significant association suggesting a dose-response effect, with heterozygotes and homozygotes exhibiting a 22% and 50% increase in motor decline, respectively.

While *MC1R’*s exact role in PD etiology is still unknown, its key role in systemic pigmentation may contribute to the observed association between *MC1R* RHC variants and accelerated PD motor decline. MC1R-mediated eumelanin synthesis may protect against oxidative stress. *MC1R* loss-of-function shifts melanin production toward pheomelanin and is associated with higher oxidative stress, abnormal skin aging, and a higher risk for melanoma.^30^ Pheomelanin synthesis also consumes glutathione, a key antioxidant implicated in PD.^31,32^ PD postmortem substantia nigra exhibit decreased eumelanin and increased pheomelanin; however, whether MC1R plays any role in neuromelanin synthesis is unknown.^9^ In mouse models of PD, *MC1R* loss-of-function leads to dopaminergic neurodegeneration while *MC1R* activation is neuroprotective.^6–8^ Mice, like other small laboratory animals, do not have detectable neuromelanin, suggesting that MC1R’s non-pigmentary functions may also play a role. MC1R activates the cAMP signaling pathway, which mediates a wide range of cellular responses, including neuroinflammation, metabolism, and DNA repair.^33^ *MC1R* RHC variants have been shown to impair cAMP coupling and may lead to decreased levels of intracellular cAMP.^34^

Our study leverages up to 13 years of longitudinal data from 802 well-characterized PPMI participants with detailed genotyping and clinical assessments to comprehensively evaluate associations between MC1R, PD progression, and phenoconversion. While our findings highlight a potentially important role for MC1R in PD progression, they should be viewed in light of several limitations. First, despite adjustment for key demographic and clinical covariates, residual confounding cannot be excluded. Unmeasured factors such as environment, comorbidities, or lifestyle may influence both *MC1R* signaling and PD progression. Second, subgroups with limited sample sizes, including participants homozygous for high-penetrance *MC1R* variants (n=17), those with rare *MC1R* variants such as D84E (n=4) and R142H (n=3), and those with prodromal PD (n=45), limited statistical power to detect genotype differences, variant-specific associations, or differences in the risk of phenoconversion. Further, our analysis was based on a binary definition of phenoconversion, whereas quantitative measures may be more informative when they become available. Nevertheless, the pronounced numerical increase in phenoconversion associated with *MC1R* RHC variants is particularly noteworthy and warrants validation in a larger cohort. Third, because MoCA and DAT-SPECT exhibited little change over follow-up, our ability to detect associations with cognitive or presynaptic dopaminergic decline was limited, and null findings should be interpreted with caution. Fourth, while sensitivity analyses suggest robustness of the motor association before and after LEDD adjustment, genetic variation in *MC1R* may influence exogenous levodopa metabolism or efficacy, and pharmacogenetic effects cannot be fully excluded. Finally, the absence of detailed skin-tone and hair-color data prevented deeper evaluation of pigmentation-related vs. non-pigmentary associations.

Here, we reveal a previously unrecognized link between *MC1R* and PD-associated motor decline. Of the 802 PD participants analyzed, 490 (61.1%) carried at least one *MC1R* RHC variant. The relatively high frequency of *MC1R* RHC variants in the PPMI cohort aligns with previous reports in other PD cohorts as well as a meta-analysis on the frequency of *MC1R* RHC variants in European populations, indicating a larger role for MC1R in PD biology than previously appreciated.^13,15–20,35^ Taken together with laboratory findings from our group and others, these results warrant validation in larger and more diverse populations, ideally incorporating quantitative neuromelanin imaging, and longitudinal biospecimens to quantify oxidative stress and peripheral melanin metabolism. Complementary laboratory work is also needed to elucidate how *MC1R* signaling influences vulnerability and progression in PD. Ultimately, this line of research may open a path toward therapeutic strategies aimed at modulating MC1R activity to alter the course of PD.

## Contributors

Conceptualization: JGS, XZ, JW, XG, MAS, MC, and XC; Data Curation: JGS and XZ; Methodology, Software, and Formal Analysis: JGS, XZ, JW, MC, and XC; Visualization: JGS and XZ; Data Validation: JGS, XZ, JW, MC, and XC; Writing – Original Draft: JGS, XZ, and XC; Writing – Review & Editing: JGS, XZ, JW, XG, MAS, MC, and XC; Supervision: XC; Funding Acquisition: XC. JGS, XZ, JW and XC have all had direct access to and have verified the underlying data. All authors have read and approved the final version of the manuscript.

## Data sharing statement

Data used in the preparation of this article is openly available to qualified researchers. Data were obtained in October 2025 from the PPMI database (www.ppmi-info.org/access-data-specimens/download-data, RRID:SCR00_6431). For up-to-date information on the study,visit www.ppmi-info.org. Code generated in this study can be found at https://github.com/JacksonGS-1/MC1R_Progression_Code.

## Declaration of interests

JGS, XZ, JW, MAS, MC, and XC declare no conflict of interest.

## Acknowledgements

This research was funded in part by Aligning Science Across Parkinson*’*s grant ASAP-000312 through the Michael J. Fox Foundation for Parkinson*’*s Research (MJFF) and by the National Institute of Health through the National Institute of Neurological Disorders and Stroke grant R01NS102735. For the purpose of open access, the author has applied a CC BY public copyright license to all Author Accepted Manuscripts arising from this submission. The authors would like to thank PPMI – a public-private partnership – funded by the Michael J. Fox Foundation for Parkinson*’*s Research and funding partners, including 4D Pharma, Abbvie, AcureX, Allergan, Amathus Therapeutics, Aligning Science Across Parkinson*’*s, AskBio, Avid Radiopharmaceuticals, BIAL, BioArctic, Biogen, Biohaven, BioLegend, BlueRock Therapeutics, Bristol-Myers Squibb, Calico Labs, Capsida Biotherapeutics, Celgene, Cerevel Therapeutics, Coave Therapeutics, DaCapo Brainscience, Denali, Edmond J. Safra Foundation, Eli Lilly, Gain Therapeutics, GE HealthCare, Genentech, GSK, Golub Capital, Handl Therapeutics, Insitro, Jazz Pharmaceuticals, Johnson & Johnson Innovative Medicine, Lundbeck, Merck, Meso Scale Discovery, Mission Therapeutics, Neurocrine Biosciences, Neuron23, Neuropore, Pfizer, Piramal, Prevail Therapeutics, Roche, Sanofi, Servier, Sun Pharma Advanced Research Company, Takeda, Teva, UCB, Vanqua Bio, Verily, Voyager Therapeutics, the Weston Family Foundation and Yumanity Therapeutics.

---

**Supplemental Table 1.** Baseline demographic and clinical characteristics for sporadic and *MC1R* PD stratified by *MC1R* variant penetrance. Data is shown as n (%), %, mean (standard deviation), or median (interquartile range). Statistical analysis comparing group characteristics was not performed. A single participant may be included in both the r Variant and R Variant groups due to the presence of compound heterozygotes. Abbreviations: PD, Parkinson*’*s disease; *MC1R*, melanocortin 1 receptor; SAA, seed amplification assay; MDS-UPDRS I and III, Movement Disorder Society Unified Parkinson*’*s Disease Rating Scale Part I and III; MoCA, Montreal Cognitive Assessment; DAT-SPECT, dopamine transporter imaging with single-photon emission computed tomography; SBR, specific binding ratio.

|  | Sporadic PD | r Variant *MC1R* PD | R Variant *MC1R* PD |
| --- | --- | --- | --- |
| N | 185 | 150 | 102 |
| *MC1R* compound heterozygotes | 0 | 22 (14.67) | 42 (41.2) |
| *MC1R* homozygotes | 0 | 13 (8.67) | 4 (3.92) |
| Age at baseline (years) | 65.2 (9.88) | 61.3 (10.1) | 61.2 (9.98) |
| Years since original diagnosis | 0.76 (1.51) | 0.63 (0.73) | 0.62 (0.74) |
| Age at PD onset | 64.4 (9.96) | 60.7 (10.1) | 60.6 (10.1) |
| Male | 112 (60.5) | 103 (68.7) | 66 (64.7) |
| Race |  |  |  |
| White | 168 (90.8) | 133 (88.7) | 96 (94.1) |
| Asian | 1 (0.54) | 7 (4.67) | 0 |
| Black | 4 (2.16) | 2 (1.33) | 1 (0.98) |
| Multiracial | 7 (3.78) | 4 (2.65) | 2 (1.97) |
| Unknown | 5 (2.70) | 4 (2.65) | 3 (2.95) |
| Hispanic or Latino ethnicity | 19 (10.3) | 4 (2.65) | 3 (2.94) |
| Relatives with PD |  |  |  |
| Parent | 43 (23.2) | 13 (8.67) | 11 (10.8) |
| Other | 46 (24.9) | 24 (16.0) | 21 (20.6) |
| α-Synuclein SAA status |  |  |  |
| Positive | 112 | 138 | 97 |
| Negative | 9 | 5 | 3 |
| Unknown | 64 | 7 | 2 |
| α-Synuclein SAA positivity | 92.6 | 96.5 | 95.1 |
| History of melanoma | 1 (0.54) | 4 (2.65) | 1 (0.98) |
| MDS-UPDRS I score at baseline | 4.41 (3.18) | 4.63 (3.45) | 4.51 (3.34) |
| MDS-UPDRS III score at baseline | 22.7 (9.6) | 20.4 (8.99) | 19.3 (8.76) |
| MoCA score at baseline | 26.8 (2.35) | 26.9 (2.75) | 27.5 (2.30) |
| DAT-SPECT SBR at baseline |  |  |  |
| Caudate | 1.93 (1.63, 2.28) | 1.99 (1.65, 2.32) | 2.02 (1.68, 2.36) |
| Putamen | 0.77 (0.63, 0.91) | 0.84 (0.67, 0.99) | 0.87 (0.65, 1.00) |

**Supplemental Table 2.** Baseline demographic and clinical characteristics for sporadic and *MC1R* PD stratified by *MC1R* genotype. Data is shown as n (%), %, mean (standard deviation), or median (interquartile range). Statistical analysis comparing group characteristics was not performed. Abbreviations: PD, Parkinson*’*s disease; *MC1R*, melanocortin 1 receptor; SAA, seed amplification assay; MDS-UPDRS I and III, Movement Disorder Society Unified Parkinson*’*s Disease Rating Scale Part I and III; MoCA, Montreal Cognitive Assessment; DAT-SPECT, dopamine transporter imaging with single-photon emission computed tomography; SBR, specific binding ratio.

|  | Sporadic PD | Single Heterozygote *MC1R* PD | Compound Heterozygote *MC1R* PD | Homozygote *MC1R* PD |
| --- | --- | --- | --- | --- |
| N | 185 | 139 | 64 | 17 |
| r variant *MC1R* PD | 0 | 83 (59.7) | 22 (34.4) | 13 (76.5) |
| R variant *MC1R* PD | 0 | 56 (40.3) | 42 (65.6) | 4 (23.5) |
| Age at baseline (years) | 65.2 (9.88) | 61.2 (10.1) | 61.3 (10.2) | 64.2 (11.3) |
| Years since original diagnosis | 0.76 (1.51) | 0.66 (0.86) | 0.60 (0.54) | 0.39 (0.28) |
| Age at PD onset | 64.4 (9.96) | 60.5 (10.1) | 60.7 (10.1) | 63.8 (11.3) |
| Male | 112 (60.5) | 97 (69.8) | 44 (68.8) | 7 (41.2) |
| Race |  |  |  |  |
| White | 168 (90.8) | 128 (92.1) | 57 (89.1) | 14 (82.4) |
| Asian | 1 (0.54) | 1 (0.72) | 3 (4.70) | 3 (17.6) |
| Black | 4 (2.16) | 3 (2.15) | 0 | 0 |
| Multiracial | 7 (3.78) | 4 (2.88) | 2 (3.10) | 0 |
| Unknown | 5 (2.70) | 3 (2.15) | 2 (3.10) | 0 |
| Hispanic or Latino ethnicity | 19 (10.3) | 1 (0.72) | 3 (4.70) | 0 |
| Relatives with PD |  |  |  |  |
| Parent | 43 (23.2) | 13 (9.35) | 7(10.9) | 1 (5.88) |
| Other | 46 (24.9) | 27 (19.4) | 12 (18.8) | 0 |
| α-Synuclein SAA status |  |  |  |  |
| Positive | 112 | 126 | 60 | 17 |
| Negative | 9 | 7 | 1 | 0 |
| Unknown | 64 | 6 | 3 | 0 |
| α-Synuclein SAA positivity | 92.6 | 94.7 | 98.4 | 100 |
| History of melanoma | 1 (0.54) | 4 | 1 | 0 |
| MDS-UPDRS I score at baseline | 4.41 (3.18) | 4.70 (3.62) | 4.48 (2.93) | 4.00 (3.22) |
| MDS-UPDRS III score at baseline | 22.7 (9.6) | 20.2 (9.15) | 19.1 (7.78) | 20.7 (10.6) |
| MoCA score at baseline | 26.8 (2.35) | 27.1 (2.60) | 27.4 (2.52) | 27.2 (2.84) |
| DAT-SPECT SBR at baseline |  |  |  |  |
| Caudate | 1.93 (1.63, 2.28) | 2.00 (1.67, 2.36) | 2.01 (1.68, 2.27) | 2.00 (1.44, 2.48) |
| Putamen | 0.77 (0.63, 0.91) | 0.85 (0.65, 1.00) | 0.87 (0.65, 0.99) | 0.84 (0.67, 0.98) |

**Supplemental Table 3.** Baseline demographic and clinical characteristics for sporadic and *MC1R* PD stratified by *MC1R* variant. Data is shown as n (%), %, mean (standard deviation), or median (interquartile range). Statistical analysis comparing group characteristics was not performed. A single participant may be included in several variant groups due to the presence of compound heterozygotes. Abbreviations: PD, Parkinson*’*s disease; *MC1R*, melanocortin 1 receptor; SAA, seed amplification assay; MDS-UPDRS I and III, Movement Disorder Society Unified Parkinson*’*s Disease Rating Scale Part I and III; MoCA, Montreal Cognitive Assessment; DAT-SPECT, dopamine transporter imaging with single-photon emission computed tomography; SBR, specific binding ratio.

|  | Sporadic PD | R163Q/P PD | V60L PD | V92M/L PD | R151S/G/C PD | R160W PD | D294N/H PD | D84E PD | R142H PD |
| --- | --- | --- | --- | --- | --- | --- | --- | --- | --- |
| N | 185 | 41 | 77 | 54 | 50 | 43 | 11 | 4 | 3 |
| *MC1R* compound heterozygotes | 0 | 17 (41.5) | 27 (35.1) | 0 | 13 (26.0) | 3 (6.98) | 2 (18.2) | 2 (50.0) | 0 |
| *MC1R* homozygotes | 0 | 4 (9.76) | 5 (6.49) | 4 (7.41) | 2 (4.00) | 2 (4.65) | 0 | 0 | 0 |
| Age at baseline (years) | 65.2 (9.88) | 61.2 (10.1) | 61.9 (11.0) | 60.4 (9.85) | 62.8 (10.9) | 62.2 (9.12) | 57.5 (7.67) | 56.8 (7.50) | 58.3 (9.29) |
| Years since original diagnosis | 0.76 (1.51) | 0.61 (0.56) | 0.70 (0.86) | 0.52 (0.53) | 0.61 (0.87) | 0.66 (0.61) | 0.58 (0.52) | 0.29 (0.14) | 0.75 (0.08) |
| Age at PD onset | 64.4 (9.96) | 60.6 (9.99) | 61.2 (10.9) | 59.9 (9.84) | 62.2 (11.1) | 61.5 (9.19) | 56.9 (7.77) | 56.5 (7.62) | 57.6 (9.21) |
| History of melanoma | 1 (0.54) | 0.00 | 2 (2.60) | 3 (5.56) | 0.00 | 1 (2.32) | 0 | 0 | 0.00 |
| Male | 112 (60.5) | 29 (70.7) | 54 (70.1) | 36 (66.67) | 29 (58.0) | 29 (67.4) | 7 (63.6) | 3 (75.0) | 3 (100) |
| Race |  |  |  |  |  |  |  |  |  |
| White | 168 (90.8) | 31 (75.6) | 72 (93.5) | 47 (87.0) | 50 (100) | 39 (90.7) | 9 (81.8) | 4 (100) | 3 (100) |
| Asian | 1 (0.54) | 6 (14.6) | 0 | 4 (7.40) | 0 | 0 | 0 | 0 | 0 |
| Black | 4 (2.16) | 0 | 0 | 2 (3.70) | 0 | 1 (2.32) | 0 | 0 | 0 |
| Multiracial | 7 (3.78) | 2 (4.90) | 3 (3.90) | 1 (1.90) | 0 | 1 (2.32) | 1 (9.10) | 0 | 0 |
| Unknown | 5 (2.70) | 2 (4.90) | 2 (2.60) | 0 | 0 | 2 (4.65) | 1 (9.10) | 0 | 0 |
| Hispanic or Latino ethnicity | 19 (10.3) | 3 (7.32) | 0 | 1 | 0 | 1 (2.32) | 1 (9.10) | 1 (25.0) | 0 |
| Relatives with PD |  |  |  |  |  |  |  |  |  |
| Parent | 43 (23.2) | 6 (14.6) | 6 (7.80) | 3 (5.56) | 8 (16.0) | 2 (4.65) | 1 (9.10) | 1 (25.0) | 0 |
| Other | 46 (24.9) | 2 (4.90) | 13 (16.90) | 12 (22.2) | 14 (28.0) | 7 (16.3) | 2 (18.2) | 1 (25.0) | 0 |
| α-Synuclein SAA status |  |  |  |  |  |  |  |  |  |
| Positive | 112 | 37 | 70 | 50 | 47 | 41 | 10 | 4 | 3 |
| Negative | 9 | 2 | 3 | 1 | 2 | 1 | 0 | 0 | 0 |
| Unknown | 64 | 2 | 4 | 3 | 1 | 1 | 1 | 0 | 0 |
| α-Synuclein SAA positivity | 92.6 | 94.9 | 95.9 | 98 | 95.9 | 97.6 | 100 | 100 | 100 |
| History of melanoma | 1 (0.54) | 0.00 | 2 (2.60) | 3 (5.56) | 0.00 | 1 (2.32) | 0 | 0 | 0.00 |
| MDS-UPDRS I score at baseline | 4.41 (3.18) | 5.80 (3.51) | 4.21 (3.04) | 4.26 (3.99) | 4.48 (3.59) | 4.49 (3.15) | 4.56 (2.13) | 7.00 (5.66) | 1.33 (1.53) |
| MDS-UPDRS III score at baseline | 22.7 (9.6) | 20.9 (9.07) | 20.6 (8.81) | 18.8 (9.10) | 19.8 (9.49) | 18.1 (7.54) | 20.7 (9.02) | 34.5 (6.36) | 15.7 (7.09) |
| MoCA score at baseline | 26.8 (2.35) | 26.4 (2.39) | 27.1 (2.84) | 27.6 (2.79) | 27.5 (2.38) | 27.3 (2.58) | 27.6 (2.40) | 29.0 (1.41) | 28.0 (2.00) |
| DAT-SPECT SBR at baseline |  |  |  |  |  |  |  |  |  |
| Caudate | 1.93 (1.63, 2.28) | 2.04 (1.72, 2.43) | 1.96 (1.53, 2.24) | 2.00 (1.74, 2.20) | 2.05 (1.75, 2.39) | 2.00 (1.67, 2.33) | 2.06 (1.49, 2.43) | 1.76 (1.52, 1.99) | 1.85 (1.60, 2.10) |
| Putamen | 0.77 (0.63, 0.91) | 0.93 (0.70, 1.06) | 0.80 (0.60, 0.97) | 0.82 (0.69, 0.99) | 0.92 (0.66, 1.05) | 0.82 (0.65, 0.92) | 0.83 (0.62, 0.92) | 0.78 (0.67, 0.89) | 0.80 (0.69, 0.99) |

**Supplemental Table 4.** Comparison of the rate of change in MDS-UPDRS I, MoCA, and DAT-SPECT SBR between participants with sporadic, *MC1R, LRRK2, GBA, MC1R + LRRK2*, and *MC1R + GBA* PD. Slopes (unit per year) were estimated from linear mixed models. Fixed effects included time ^*^ genetic group, baseline MDS-UPDRS III, baseline age, years since original diagnosis to baseline, sex, race, ethnicity, and LEDD. Participant-level random intercepts and slopes with unstructured covariance, heteroscedastic by genetic group. % difference is relative to the direction of change of each assessment. *MC1R* PD, *LRRK2* PD, and GBA PD were compared to sporadic PD in a single model while *MC1R*+*LRRK2* PD and *MC1R*+GBA PD were compared to *LRRK2* PD and GBA PD in separate models. Stratification by penetrance, genotype, and variant was performed in separate models using sporadic PD as the reference group. A single participant may be included in multiple groups due to the presence of compound heterozygotes. N refers to the comparison group | reference group. Abbreviations: PD, Parkinson*’*s disease; *MC1R*, melanocortin 1 receptor; MDS-UPDRS I and III, Movement Disorder Society Unified Parkinson*’*s Disease Rating Scale Part I and III; MoCA, Montreal Cognitive Assessment; DAT-SPECT, dopamine transporter imaging with single-photon emission computed tomography; SBR, specific binding ratio.

| Group | N | MDS-UPDRS I |  |  | MoCA |  |  | Caudate SBR |  |  | Putamen SBR |  |  |
| --- | --- | --- | --- | --- | --- | --- | --- | --- | --- | --- | --- | --- | --- |
| | | Slope, $\beta$ (95% CI) | % diff | p-value | Slope, $\beta$ (95% CI) | % diff | P-value | Slope, $\beta$ (95% CI) | % diff | p-value | Slope, $\beta$ (95% CI) | % diff | p-value |
| Sporadic PD | 185 | 0.42 (0.34, 0.50) |  |  | -0.23 (-0.32, -0.13) |  |  | -0.13 (-0.15, -0.11) |  |  | -0.06 (-0.07, -0.06) |  |  |
| *MC1R* PD vs. Sporadic PD | 220 \| 185 | 0.52, 0.10 (-0.01, 0.20) | 24% | 0.070 | -0.21, 0.02 (-0.09, 0.14) | -10% | 0.690 | -0.13, 0.00 (-0.02, 0.03) | 0% | 0.908 | -0.06, 0.00 (-0.01, 0.01) | 0% | 0.827 |
| r variant *MC1R* PD | 150 \| 185 | 0.49, 0.07 (-0.05, 0.19) | 17% | 0.225 | -0.30, -0.07 (-0.22, 0.07) | 30% | 0.324 | -0.13, 0.00 (-0.03, 0.02) | 0% | 0.868 | -0.06, 0.00 (-0.02, 0.01) | 0% | 0.682 |
| R variant *MC1R* PD | 102 \| 185 | 0.55, 0.13 (0.00, 0.16) | 31% | 0.055 | -0.13, 0.10 (-0.03, 0.23) | -30% | 0.143 | -0.12, 0.01 (-0.02, 0.04) | -8% | 0.707 | -0.06, 0.00 (-0.02, 0.02) | 0% | 0.915 |
| Single Heterozygote *MC1R* PD | 139 \| 185 | 0.49, 0.07 (-0.04, 0.19) | 17% | 0.202 | -0.19, 0.04 (-0.09, 0.16) | -17% | 0.575 | -0.13, 0.00 (-0.03, 0.03) | 0% | 0.925 | -0.06, 0.00 (-0.01, 0.02) | 0% | 0.780 |
| Compound Heterozygote *MC1R* PD | 64 \| 185 | 0.50, 0.08 (-0.08, 0.23) | 19% | 0.352 | -0.18, 0.05 (-0.13, 0.23) | -22% | 0.596 | -0.12, 0.01 (-0.02, 0.04) | -8% | 0.620 | -0.07, -0.01 (-0.03, 0.01) | 17% | 0.315 |
| Homozygote *MC1R* PD | 17 \| 185 | 0.79, 0.37 (0.12, 0.62) | 88% | **0.003** | -0.40, -0.17 (-0.46, 0.12) | 74% | 0.251 | -0.13, 0.00 (-0.06, 0.05) | 0% | 0.942 | -0.05, 0.01 (-0.02, 0.03) | -17% | 0.668 |
| R163Q/P PD | 41 \| 185 | 0.52, 0.10 (-0.08, 0.28) | 24% | 0.273 | -0.08, 0.15 (-0.05, 0.35) | -65% | 0.131 | -0.13, 0.00 (-0.03, 0.04) | 0% | 0.821 | -0.06, 0.00 (-0.02, 0.02) | 0% | 0.665 |
| V60L PD | 77 \| 185 | 0.47, 0.05 (-0.09, 0.19) | 12% | 0.487 | -0.13, 0.10 (-0.06, 0.26) | -43% | 0.224 | -0.12, 0.01 (-0.03, 0.04) | -8% | 0.745 | -0.06, 0.00 (-0.02, 0.02) | 0% | 0.972 |
| V92M/L PD | 54 \| 185 | 0.47, 0.05 (-0.15, 0.24) | 12% | 0.650 | -0.19, 0.04 (-0.18, 0.26) | -17% | 0.741 | -0.16, -0.03 (-0.08, 0.01) | 23% | 0.183 | -0.06, 0.00 (-0.03, 0.02) | 0% | 0.771 |
| R151S/G/C PD | 50 \| 185 | 0.53, 0.11 (-0.04, 0.26) | 26% | 0.157 | -0.36, -0.13 (-0.30, 0.04) | -57% | 0.140 | -0.12, 0.01 (-0.02, 0.05) | -8% | 0.386 | -0.07, -0.01 (-0.03, 0.02) | 17% | 0.627 |
| R160W PD | 43 \| 185 | 0.45, 0.03 (-0.14, 0.20) | 7% | 0.726 | -0.25, -0.02 (-0.20, 0.17) | 9% | 0.852 | -0.12, 0.01 (-0.03, 0.06) | -8% | 0.566 | -0.05, 0.01 (-0.01, 0.03) | -17% | 0.498 |
| D294N/H PD | 11 \| 185 | 0.57, 0.15 (-0.22, 0.52) | 36% | 0.431 | -0.13, 0.10 (-0.32, 0.53) | -43% | 0.627 | -0.15, -0.02 (-0.10, 0.06) | 15% | 0.681 | -0.06, 0.00 (-0.05, 0.04) | 0% | 0.915 |
| D84E PD | 4 \| 185 | 0.59, 0.17 (-0.63, 0.96) | 40% | 0.678 | 0.15, 0.38 (-0.46, 1.22) | -165% | 0.374 | -0.07, 0.06 (-0.04, 0.16) | -46% | 0.241 | -0.07, -0.01 (-0.09, 0.08) | 17% | 0.878 |
| R142H PD | 3 \| 185 | 1.02, 0.60 (0.03, 1.17) | 143% | **0.039** | -0.20, 0.03 (-0.57, 0.62) | -13% | 0.932 | -0.14, -0.01 (-0.12, 0.10) | 8% | 0.830 | -0.02, 0.04 (-0.04, 0.11) | -67% | 0.359 |
| *LRRK2* PD vs. Sporadic PD | 84 \| 185 | 0.26, -0.16 (-0.30, -0.01) | -38% | **0.040** | -0.04, 0.19 (0.02, 0.36) | -83% | **0.027** | -0.12, 0.01 (-0.03, 0.05) | -8% | 0.647 | -0.06, 0.00 (-0.02, 0.02) | 0% | 0.812 |
| *GBA* PD vs. Sporadic PD | 43 \| 185 | 0.51, 0.09 (-0.13, 0.31) | 21% | 0.413 | -0.20, 0.03 (-0.20, 0.26) | -13% | 0.803 | -0.15, -0.02 (-0.08, 0.04) | 15% | 0.509 | -0.07, -0.01 (-0.04, 0.01) | 14% | 0.319 |
| *MC1R* + *LRRK2* PD vs. Sporadic PD | 187 \| 185 | 0.32, -0.10 (-0.23, 0.03) | -24% | 0.138 | -0.13, 0.10 (-0.04, 0.24) | -43% | 0.160 | -0.11, 0.02 (-0.01, 0.05) | -15% | 0.124 | -0.05, 0.01 (-0.01, 0.03) | -14% | 0.183 |
| *MC1R* + *GBA* PD vs. Sporadic PD | 83 \| 185 | 0.43, 0.01 (-0.15, 0.17) | 2% | 0.894 | -0.26, -0.03 (-0.21, 0.15) | 13% | 0.725 | -0.13, 0.00 (-0.04, 0.04) | 0% | 0.882 | -0.05, 0.01 (-0.02, 0.03) | -14% | 0.577 |
| *MC1R* + *LRRK2* PD vs. *LRRK2* PD | 187 \| 84 | 0.32, 0.06 (-0.09, 0.21) | 23% | 0.429 | -0.13, -0.09 (-0.26, 0.08) | 225% | 0.312 | -0.11, 0.01 (-0.03, 0.06) | -8% | 0.538 | -0.05, 0.01 (-0.02, 0.03) | -14% | 0.552 |
| *MC1R* + *GBA* PD vs. *GBA* PD | 83 \| 43 | 0.43, -0.08 (-0.33, 0.16) | -16% | 0.515 | -0.26, -0.06 (-0.32, 0.19) | 30% | 0.615 | -0.13, 0.02 (-0.05, 0.09) | -13% | 0.531 | -0.05, 0.02 (-0.02, 0.05) | -28% | 0.308 |

**Supplemental Table 5.** Demographic characteristics for PPMI participants with sporadic and *MC1R* prodromal PD. Data is shown as n (%) or mean (standard deviation). Statistical analysis comparing group characteristics was not performed. A single participant may be included in several carrier groups due to the presence of compound heterozygotes. Abbreviations: PD, Parkinson*’*s disease; *MC1R*, melanocortin 1 receptor

|  | Sporadic<br>Prodromal PD | *MC1R*<br>Prodromal PD |
| --- | --- | --- |
| N | 16 | 29 |
| R163Q/P carriers | ---- | 6 (20.7) |
| V60L carriers | ---- | 7 (24.1) |
| V92M/L carriers | ---- | 6 (20.7) |
| R151S/G/C carriers | ---- | 6 (20.7) |
| R160W carriers | ---- | 7 (24.1) |
| D294N/H carriers | ---- | 0 |
| D84E carriers | ---- | 0 |
| R142H carriers | ---- | 3 (10.3) |
| *MC1R* compound heterozygotes | ---- | 6 (20.7) |
| *MC1R* homozygotes | ---- | 2 (6.89) |
| Phenoconverters | 4 (25.0) | 18 (62.1) |
| Age at baseline (years) | 68.8 (6.20) | 69.6 (5.82) |
| Average follow-up (years) | 6.14 (3.16) | 8.31 (2.72) |
| Male | 16 (100) | 22 (75.9) |
| Race |  |  |
| White | 14 (87.5) | 27 (93.1) |
| Black | 1 (6.25) | 1 (3.45) |
| Unknown | 1 (6.25) | 1 (3.45) |
| Hispanic or Latino ethnicity | 16 (100) | 22 (75.9) |
| Relatives with PD |  |  |
| Parent | 2 (12.5) | 1 (3.45) |
| Other | 0 | 2 (6.90) |
| History of melanoma | 0 | 0 |

## Notes

### Competing Interest Statement

The authors have declared no competing interest.

### Author Declarations

Data used in the preparation of this article is openly available to qualified researchers. Data were obtained in October 2025 from the PPMI database (www.ppmi-info.org/access-data-specimens/download-data, RRID:SCR00_6431).

